# Using Quality Improvement Tools to Reduce Macerated Stillbirth by 50% in the Volta Region of Ghana in a Two Years Period

**DOI:** 10.1101/2024.10.31.24316366

**Authors:** Gideon Danso, Richard Okyere Boadu

## Abstract

Stillbirth rate is an indirect measure of the quality of management of pregnancy, labour and delivery. The loss of a baby due to stillbirth does not only affect pregnant women but also takes a great toll on family’s health and wellbeing. In Ghana, stillbirths account for a large proportion of perinatal mortalities. We used small multiple graphs to compare the rate of stillbirth in the Volta region from (2014-2016) to the national target for stillbirth.

Inferences from the graph shows that there was a gap of 10.4 per 1,000 live births between the national target of 12 per 1,000 live births. Fresh still birth also had a mean of 37.2 per 1000 live births showing a gap of 25.2 between the national target. The gap between the national target and macerated still birth is 50.8. This shows significant change in the stillbirth rate in the region and requires attention. In view of this, this study aims to reduce stillbirth (macerated) by 50% in two years period.

## Introduction

Stillbirths are a reflection of the quality of care given to women and newborns during antenatal and delivery (GHS annual report, 2016) is defined by the Ghana Health Service as a baby born with no signs of life at or after twenty-eight (28) weeks of gestation. The loss of a baby due to stillbirth takes a great toll on family’s health and wellbeing, still birth remains a sad reality for many families (CDC, 2020). Maternal and Child health remains at the core of global health priorities transcending the Millennium Development Goals into the current era of Sustainable Development Goals. Most low and middle-income countries including Ghana are yet to achieve the required levels of reduction in child and maternal mortality (Id et al., 2020).Most stillbirths in LMIC are considered to be preventable through provision of quality care for all mothers and babies.(Aminu et al., 2019). Investing more in maternal care and the care of small and sick newborns during labour and childbirth is perhaps the most important step in saving the lives of mothers and newborns (EMEM guide, 2017). The two types of still birth are fresh and macerated stillbirth. Fresh stillbirth is the intrauterine death of a fetus during labor or delivery while macerated stillbirth is the intrauterine death of a fetus more than twelve hours of delivery with signs of maceration. Macerated stillbirth has many causes ranging from maternal diseases and infections, fetal complications lick problem with umbilical cord and emotional and psychological trauma or stress.

**Figure 1:**
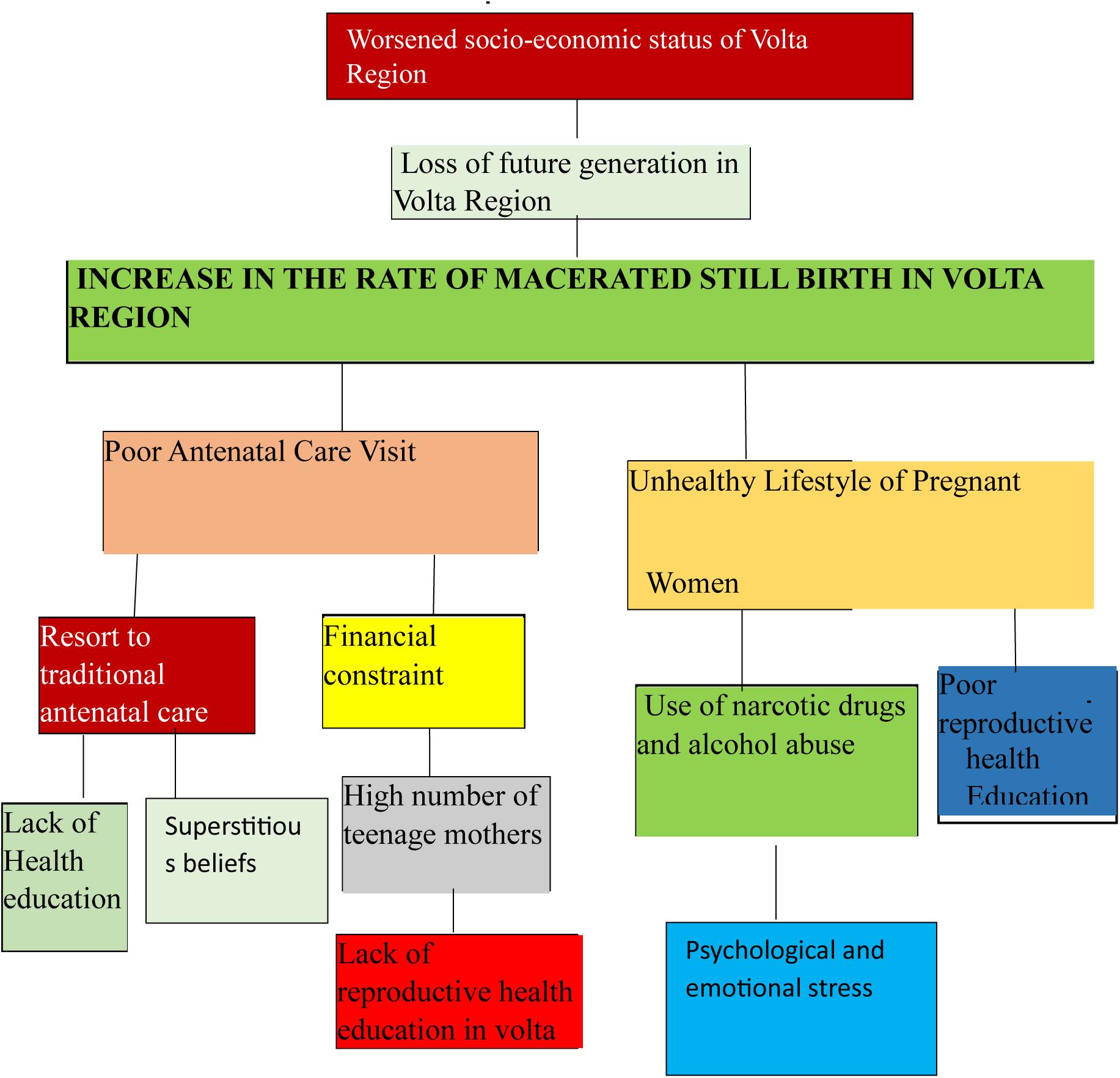
Problem Analysis (Using the Problem Tree and Five Whys)

### Problem Analysis Using the Pareto Principle

Pareto analysis is a simple technique using a bar graph to select the ideas for change that will have the greatest overall effect on the problem we want to solve and improve. In using the Pareto Analysis, we identified the causes that have the greatest impact on our main problem, which assisted in determining where to focus our improvement efforts. The principle sets out that the majority of results in any given situation will usually be determined by a small number of causes (the ‘law of the vital few’). Put simply, roughly 80% of results are determined by 20% of the causes (also called the 80:20 rule).

### How We Constructed Our Pareto Chart

We identified the problem and possible causes revealed by the five whys and problem tree QI tools.

We collected data on the causes we identified by frequency

A table was created and the causes were ranked from largest to smallest.

We constructed a bar graph with the causes along the horizontal axis and the frequency on the vertical axis.

We made a cumulative frequency graph by stacking the bars on top of each other in order of frequency, with the most common cause at the bottom and the least common at the top.

We then observed which causes accounted for the majority of the problem by looking for those that make up approximately 80% of the total number of causes counted.

#### PARETO ANALYSIS DATA

Table 1 above shows the collated data for the Pareto analysis. Out of a total of 100%, the ‘total number of occasions column’ shows the percentage occurrence of a cause. The percentage occurrence is assumed for the purpose of the study.

**Table 1:** Pareto analysis data.

| Cause | Total Number of Occasions |
| --- | --- |
| Lack of health reproductive health education in Volta region | 30 |
| Inconsistent visit to antenatal care | 20 |
| Psychological and emotional breakdown | 18 |
| Unhealthy lifestyle of pregnant women | 12 |
| Long Distance to health facility | 9 |
| Financial Constraints | 6 |
| Lack of in-depth t In-service training | 5 |

*Source: QI 2022 project*

**Figure 1.1:**
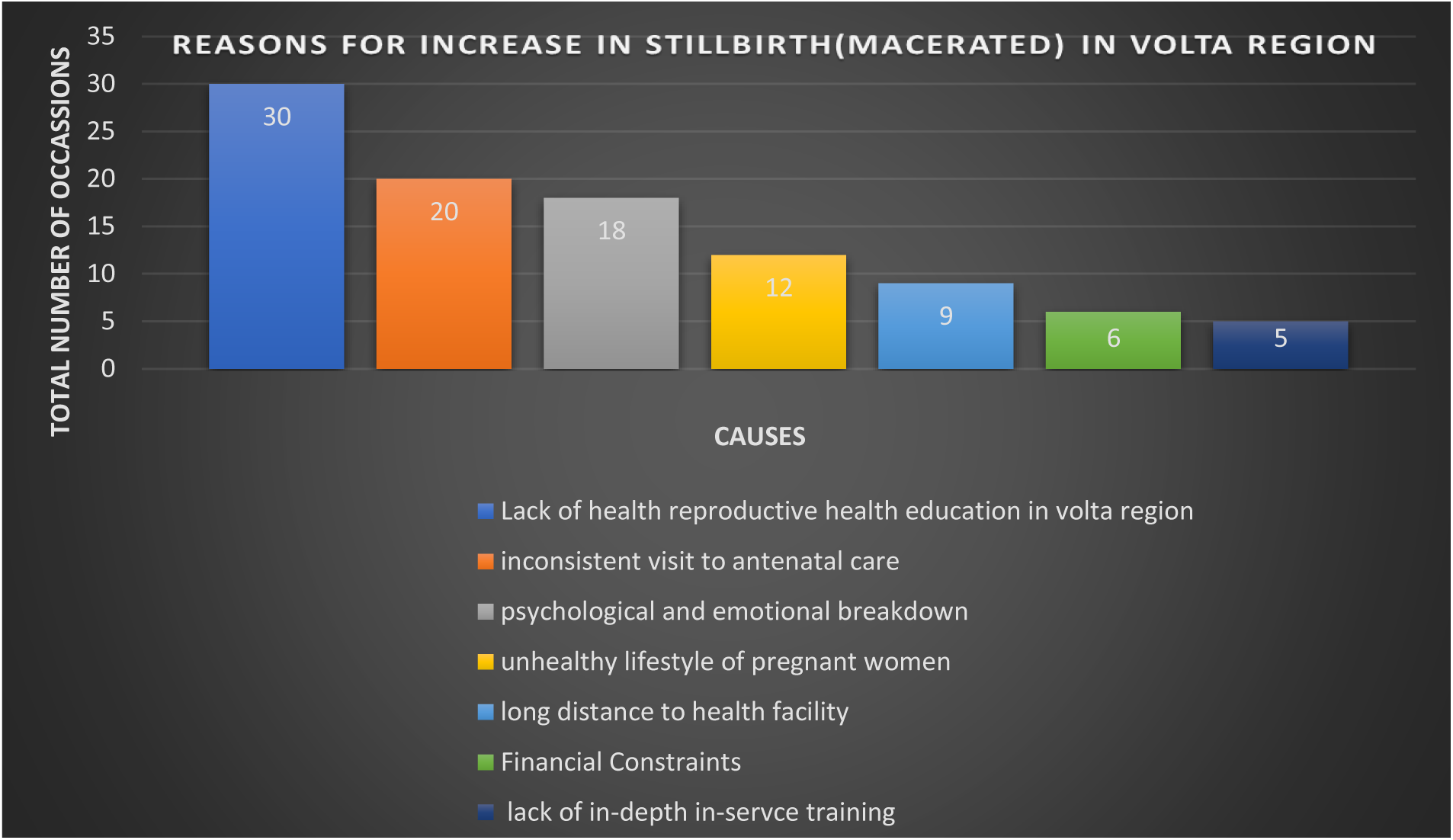
A Bar Chart Showing Reasons for Increase in macerated Still birth rate in the eight facilities in the Volta Region

**Figure 1.2:**
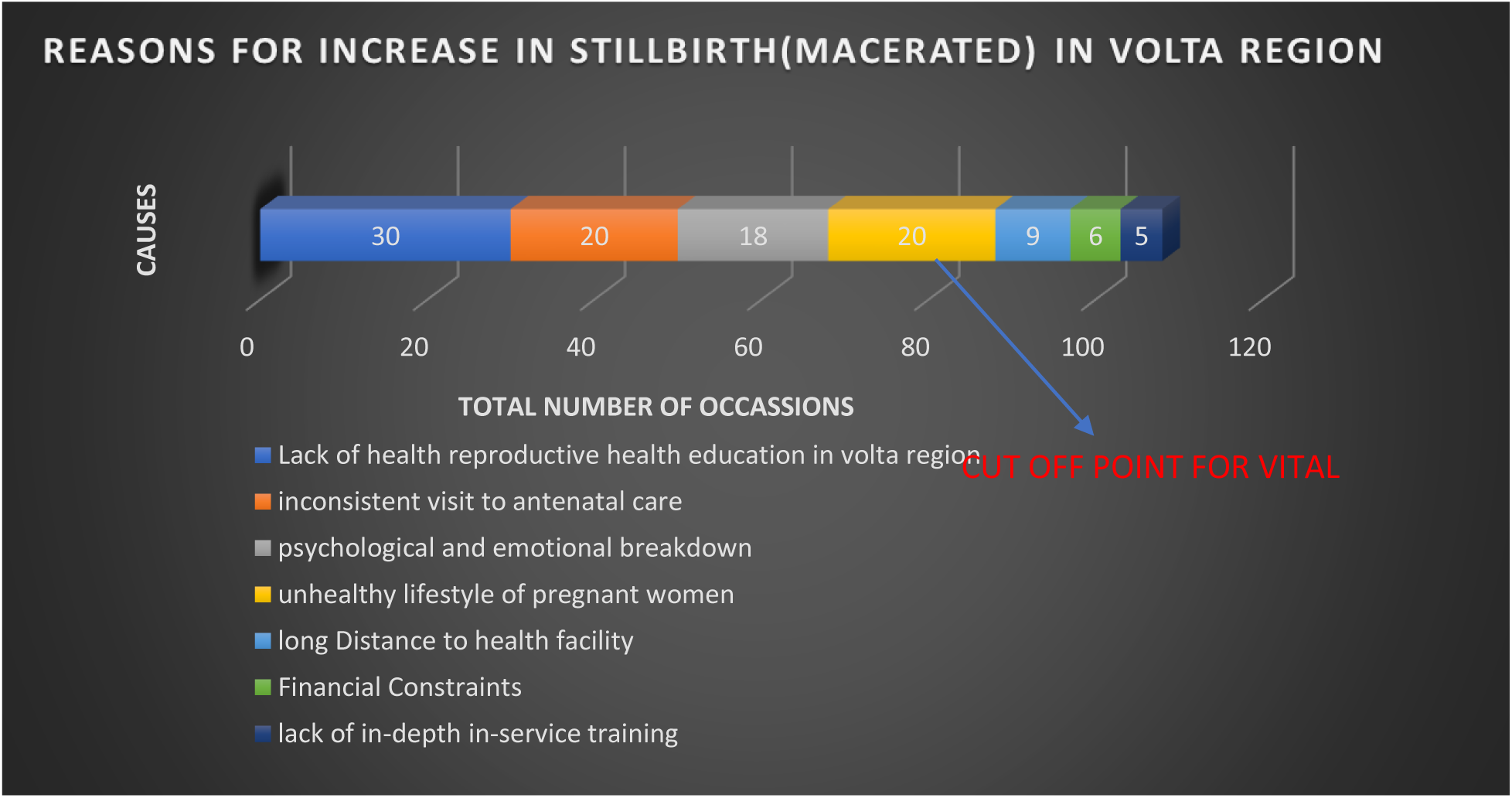
Cumulative Bar Chart Showing the Reasons for Increase in macerated Stillbirth Rate in Volta Region

By using the Pareto analysis, we have realized that only four out of the seven root causes are responsible for 80% of the problem. These four causes are Lack of reproductive health education in Volta Region, inconsistent visit to ANC, psychological and emotional stress and unhealthy lifestyle of pregnant women. By focusing much effort and resources solving these four problems, we would be actually solving 80% of the problem.

### Using the Brainstorming and Affinity Grouping QI Tools to Generate Ideas and Solutions

Brainstorming is an idea generation tool designed to produce a large number of ideas through the interaction of a group of people. An affinity diagram is a way to organize ideas into coherent patterns or themes, based on their natural relationship. It encourages people to think inventively and make non-traditional connections between ideas. It is especially useful at stages where you have generated a large volume of ideas.

Our team brainstormed using brainstorming as a QI tool and came out with strategic solutions to the various causes of the high increase in the rate of macerated stillbirth in the Volta Region. The ideas and solutions generated were organized into coherent themes based on their natural relationships. The organization of ideas was based on the affinity grouping. The diagram is shown below.

The ideas generated using the brainstorming tool were displayed by posting them in random order on a table.

Without talking, we sorted the ideas into related groups:

First, we looked for two ideas that seem to be related in some way and placed them in a separate group.

We continued to sort through the notes, establishing new groups or adding notes to existing groups.

We kept doing this till the process was complete. That is, when all the notes had been assigned to a group.

We created a title for each group that captures the relationship between the ideas contained in the notes.

Once the sorting process had been completed, we reviewed the results together.

The final solutions were decided and their details were put in a table. The table is shown below:

| CAUSES OF PROBLEM | SUGGESTED SOLUTIONS | SOLUTION DETAILS |
| --- | --- | --- |
| Inconsistent visit to ANC, financial constraint, long distance to health facility | Make mobile antenatal and postnatal services available to the catchment areas of the Volta region.<br>Pool resources from grants, donations and tokens. | Make a visit to the various areas where clients come from and organize antenatal and postnatal services to them to solve the distance barrier between them and the facility. Seek sponsorships from NGO's to be able to reach out to the catchment areas to provide mobile services and also purchase some medical supplies for them. |
| Lack of reproductive education, Unhealthy lifestyle of pregnant women, emotional and psychological stress | Organize reproductive health education and educative talk shows for both men and women on the healthy diet and lifestyle for a safe pregnancy, and infections that cause macerated stillbirth and also how to manage stress during pregnancy. | This Organize reproductive health education and educative talk show shall seek to engage the people especially the target population (women) and their husbands about healthy lifestyles and diet plan for safe delivery, and the management of stress.<br>The men will be educated on how to support their wives during pregnancy period and the changes to expect in the women. |
| Lack of in-depth in-service training | Conduct an intensive In-Service Training for all Frontline Workers (Nurses, midwives and doctors) of the maternal unit in selected eight hospitals in the Volta region | Organize training sessions for frontline ANC and delivery staff on current knowledge on delivery of all forms and the right ways to handle a pregnant woman during check-up. Senior nursing officers will be well equipped to take junior nurses through pregnancy school education |

*Source: QI 2022 project*

### Main objective

The goal of this study is to reduce macerated stillbirth by 50% in the Volta Region in two years’ time period.

### Specific objectives

Educating women on reproductive health

Educate women and men on healthy lifestyle and psychological and emotional stress management. Providing mobile services to pregnant women.

#### PLAN

We would develop a plan to test the change, to find out Who, What? When? Where? What data need to be collected?)

#### DO

1. We carry out the test.
2. We then document problems and unexpected observations.
3. Begin analysis of the data.

#### STUDY

Complete the analysis of the data. •

Summarize and reflect on what was learned.

#### ACT

1. Adopt, adapt or abandon the plan •
2. Determine what modifications should be made. •
3. Prepare a plan for the next test. •
4. Develop a plan to test the intervention made

**Figure 1.2.**
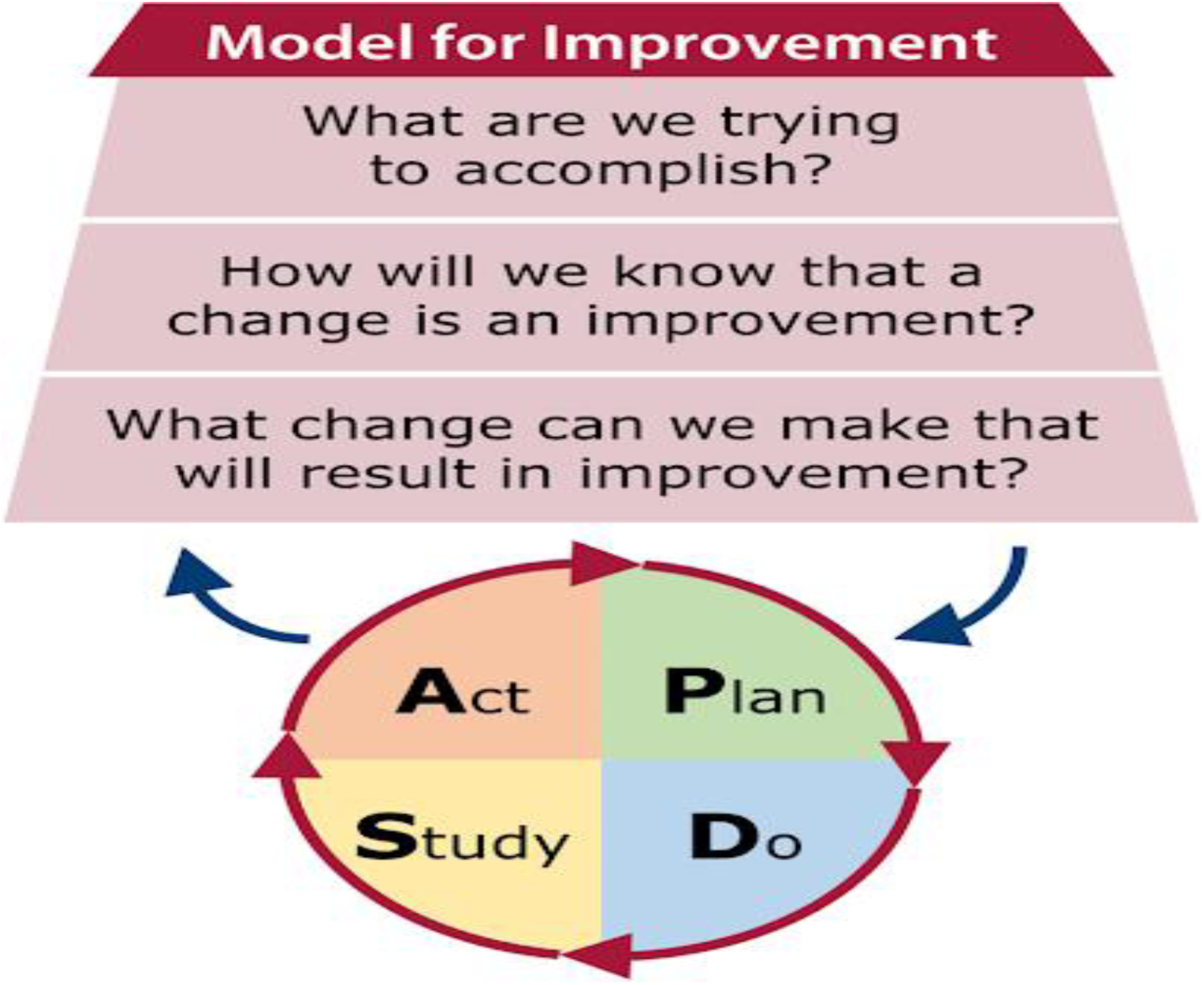
PDSA Model for Improvement

## PHASE 1: PLANNING

### Introduction

This chapter contains run diagrams for institutional stillbirth rates, problem tree analysis, objective tree analysis, key stakeholders involved in the project, project scheduling, project budget, risk factors which may be encountered in the project. We would develop a plan to test the change, to find out Who, What? When? Where? What data need to be collected?

### Dashboard for Institutional Still Birth Rate in Volta Region (2014-2018)

**Figure 1.3.**
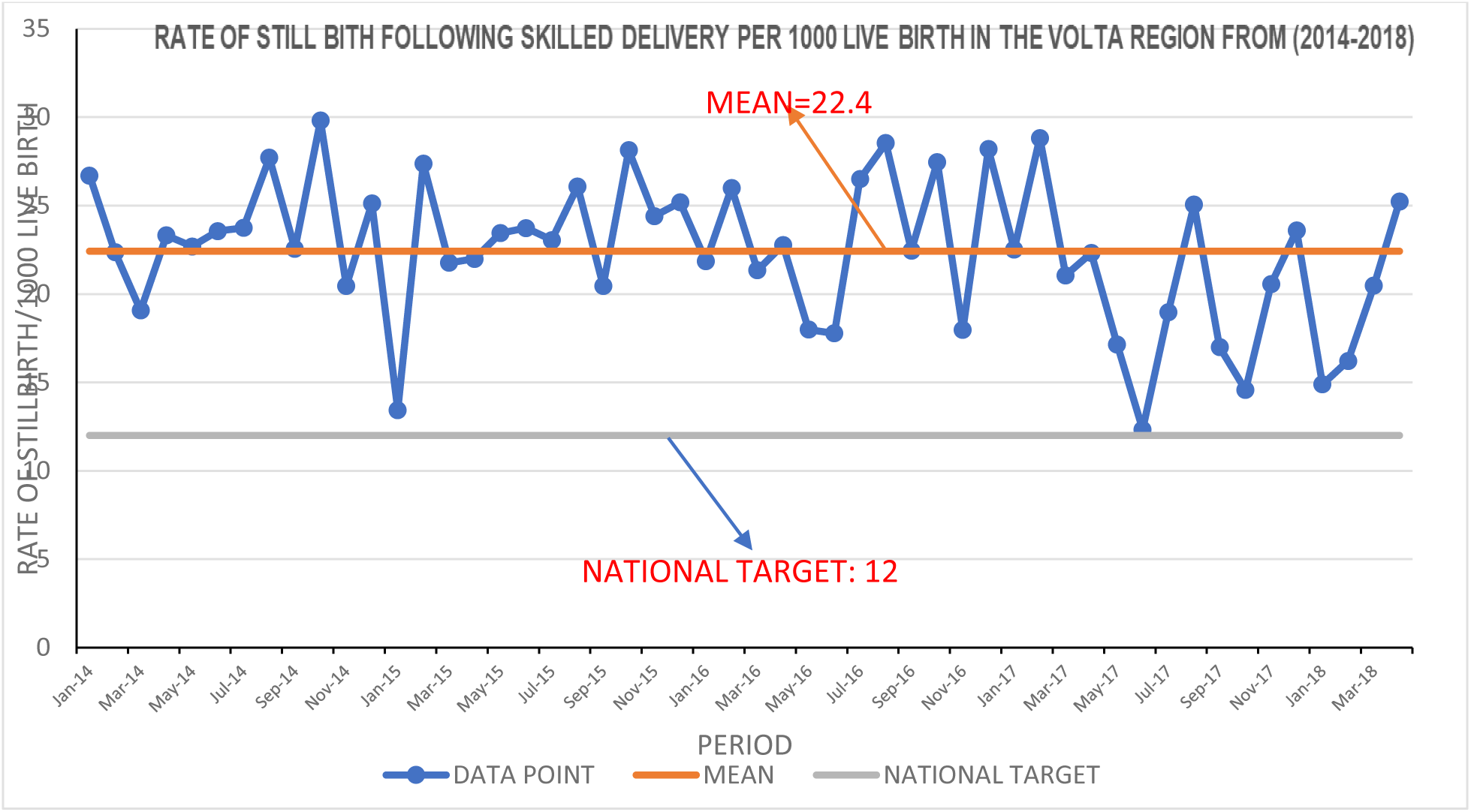

**Figure 1.4.**
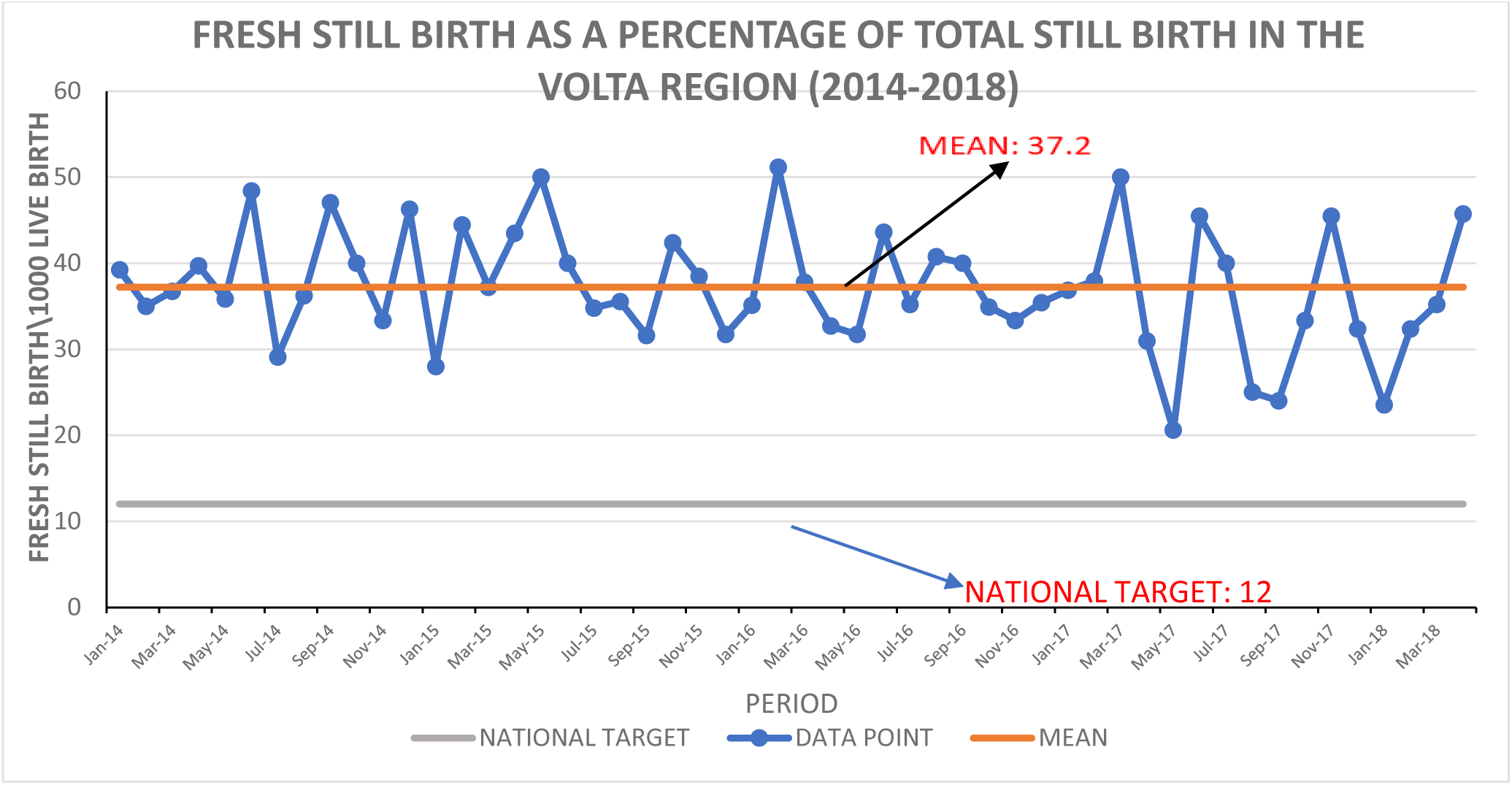

**Figure 1.5.**
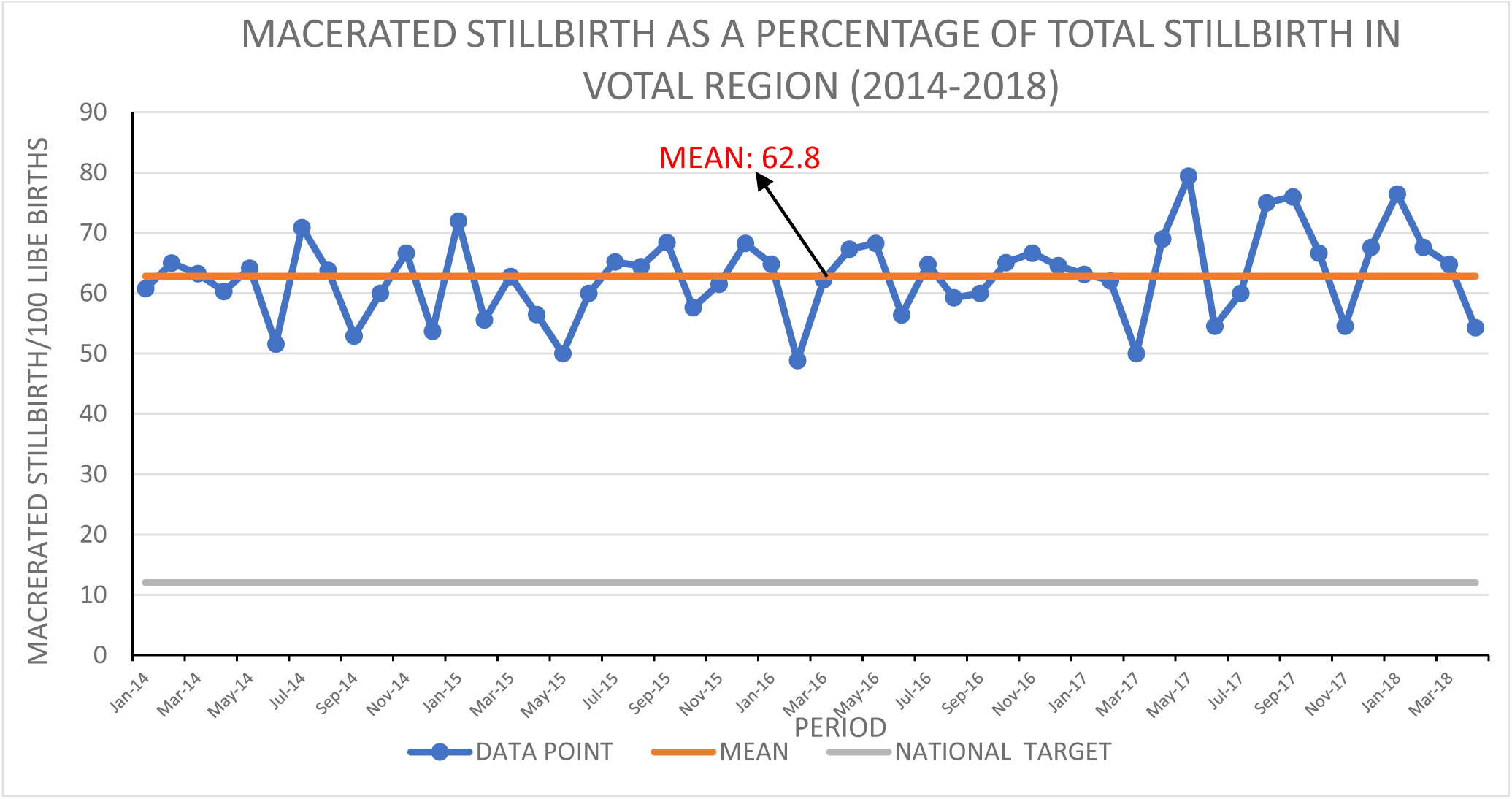

### Discussions on institutional rate of stilbirth diagrams

Figure 1.3 above, shows the rate of stillbirth following skilled delivery in the volta region from 2014 to 2018. From the graph, the mean for the rate of still birth in the volta region from 2014 to 2018 was 22.4/1000 livebirth. This means that out of 1000 livebirths, the rate of newborns who died during or before delivery was 22.4 which is above the sustainable development goal’s national target of 12/1000 livebirths. Inferences from figure 1.3 shows a gap of 10.4/1000 between the national target and the mean from 2014-2018. The highest rate was recorded in October 2014 and the lowest recorded in June 2017. Aslo, figure 1.4 above, shows the result for the fresh still birth as a percentage of the total stillbirth in the volta region from 2014 to 2018. From the graph, the mean for fresh stillbirth as a percatage of the total stillbrith was 37.2 which suggests that out the total number of babies who died in the eight (8)selected facilities in the volta region, 37.2% of them were fresh (ie, occurred during delivery). This result is far above the sustainable development goal’s national target of of 12/1000 livebirths. Inferences from figure 1.4 shows a gap of 25.2 between the national target and the mean from 2014-2018 in the volta region. The hihest percentage was recorded in February 2016 and the lowest percentage in May 2017.Finally, figure 1.5 above, shows the result for macerated stillbirtha as percentage of the total stillbirth in the volta region from 2014-2018. From the graph, the mean for fresh still birth as a percentage of the total stillbirth from 2014-2018 was 62.8 which suggest that between the years of 2014 to 2018 in the eight seleted facilities in the volta region, 62.6% of the total number of babies who died were macerated (ie, occurred before delivery). This result greatly exceeds the sustainable development goal’s national targetof 12/1000 livebirths. Inferences from the grapg suggest a huge gap of 50.8 between the mean of macerated still bith for volta region between the years odf 2014 to 2018 and the national target. The highest percentage was recorded in May 2017 and the lowest percentage in February 2016.

Looking at the three inferences drawn from figure 1,3, 1.4 and 1.5 above, it is clear that the most problematic indicator in the volta region from 2014 to 2018 was macerated stillbirth. The gap between the mean of macerated stillbirth (50,8) and the national target (12/1000) was twice the gap between the mean of mean of fresh stillbirth (25.2) and the national target (12/1000).

In view of this infernces, the group identified macerated stillbirth as the most problematic indicator and hence decided to develop strategies using PDSA QI tool to reduce the rate of macerated stillbirth in the volta region by 50% in two years period.

### Dashbord for Institutional Neonatal Death Rate and Number of Babies Referred in the Volta Region (2014-2018)

**Figure 1.6.**
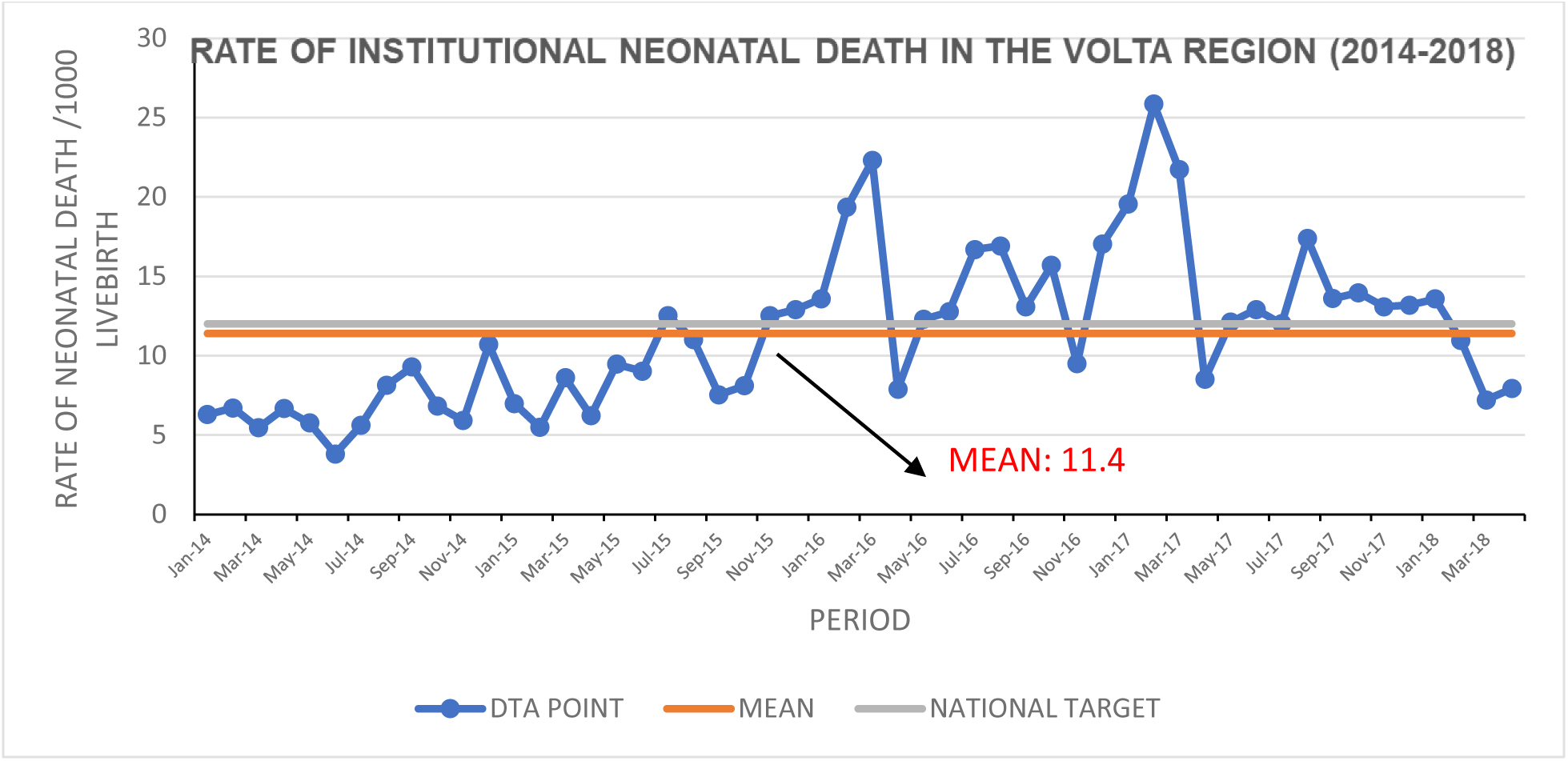

**Figure 1.7.**
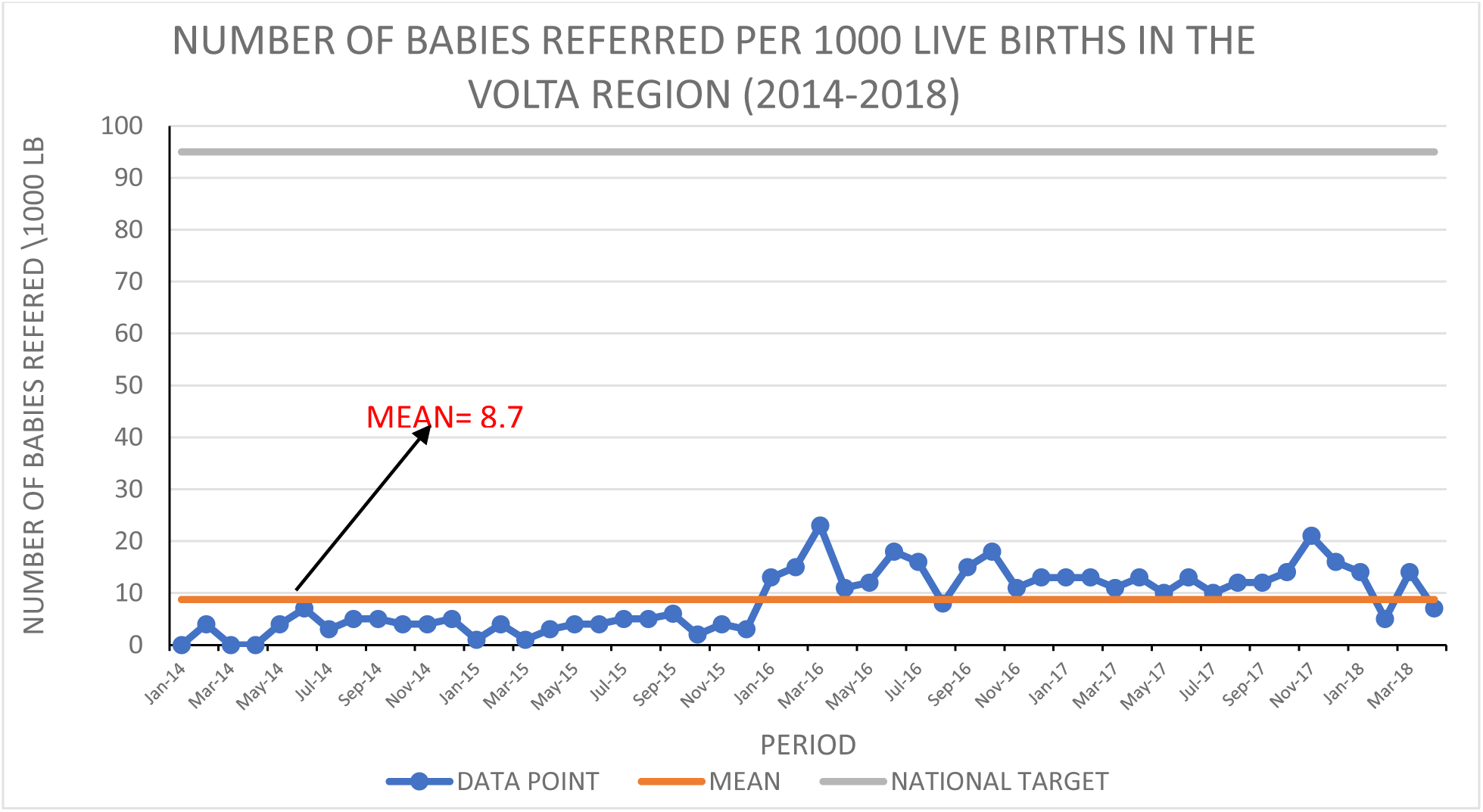

### Discussion on Institutional Neonatal Run Charts

Figure 1.6 above, shows the rate of institutional neonatal death in the Volta Region from 2014 to 2018. From the graph, it can be seen that the mean for institutional neonatal death in the Volta Region from 2014 to 2018 was 11.4 which is below the national target of 12/1000 live birth. If we are to use the national target as of 2021 which was 8.0/1000 live births, the gap between the mean of the institutional neonatal death in the Volta Region from 2014-2018 and the national target will be 3.4. This is not on the bad side as compared to the gap between the national target for macerated stillbirth (12/1000 live births) and the mean in the Volta Region from 2014 to 2018 which is 50.8. Also, figure 1.7 above shows the number of babies referred in the Volta Region between the years of 2014 to 2016. From the graph the mean for babies referred in the stated period was 8.7. This means that between the years of 2104 to 2018 in the Volta Region only 8 babies were referred from the eight (8) selected facilities.

### The Fishbone Diagram Fishbone Diagram of Major Factors Causing High Macerated Stillbirth

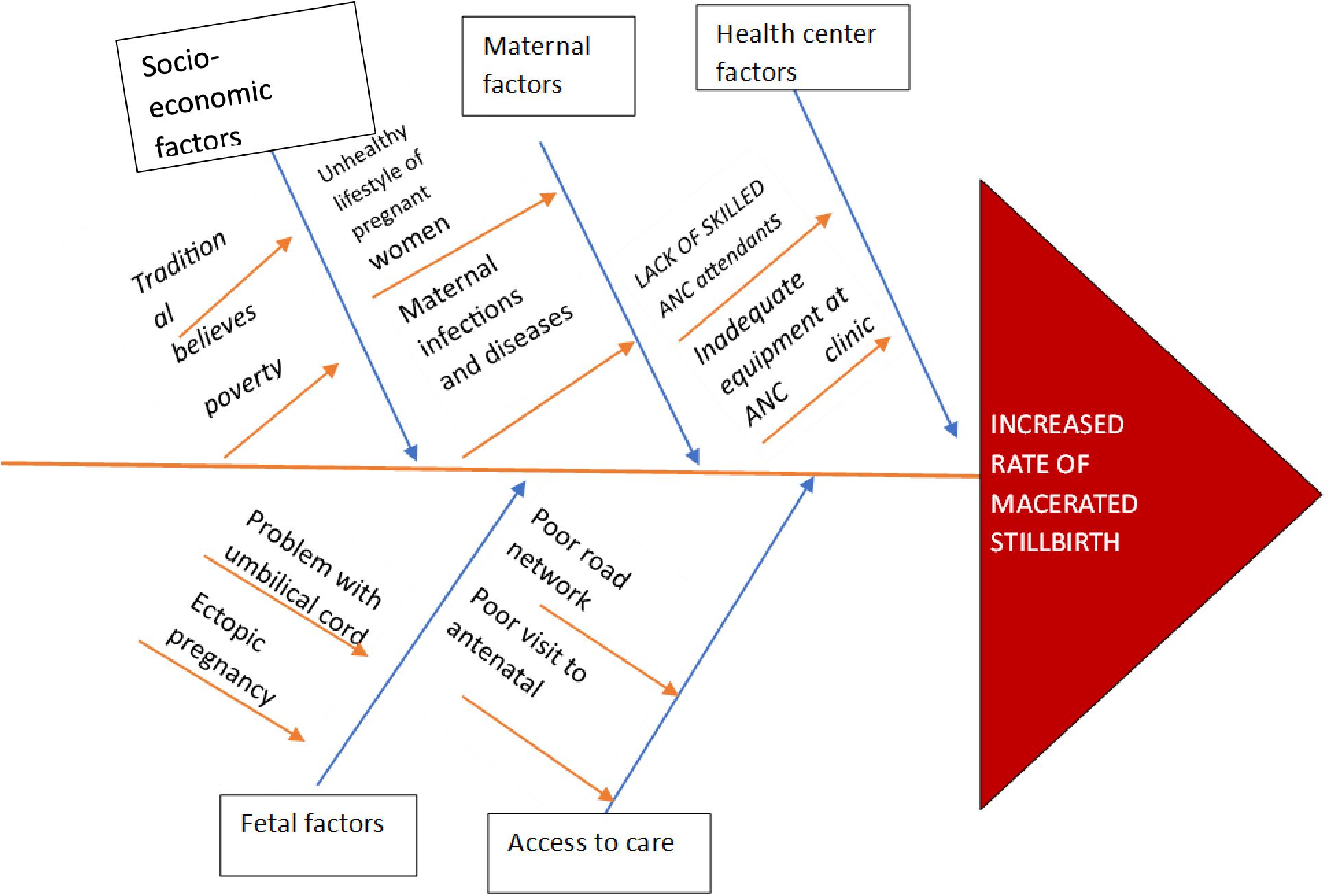

### Problem and Objective Tree

**Table 3:** Table showing the problem and objective.

| PROBLEMS | OBJECTIVES |
| --- | --- |
| **Lack of reproductive health education** | Educating women on reproductive health and some infections that cause premature death of babies. |
| **Unhealthy lifestyle of pregnant women** | Educate women on healthy lifestyle for safe pregnancy |
| **Psychological and emotional stress** | Educate women and men on psychological and emotional stress management |
| **Long distance to health facility** | Provide mobile ANC services to pregnant women |

*Source: QI 2022 project*

### Log frame

Viewing the quality improvement process as a project we used a log frame to come up with our goals, objectives, outputs and outcomes for each solution. The result is displayed in the table below:

| NARRATIVE SUMMARY | INDICATORS | DATA SOURCES | ASSUMPTIONS |
| --- | --- | --- | --- |
| **Goal: Reduce the rate of macerated Stillbirth in the eight (8) selected facilities in the Volta Region by 75% in two years period.** | The eight (selected facilities in the Volta Region) | DHIMS 2<br>Statement of inpatient in the maternity unit from the selected facilities in the Volta Region. | Quality of data is sufficient |
| **Objective 1: Educating women especially pregnant women who visit ANC in the selected facilities on reproductive health and some infections that can cause premature death of babies.** | Number of pregnant women who had macerated stillbirth due to infections | Reproductive and Child Health report (RCH)<br>Consulting room register | Mothers at Catchment Areas respond positively to call to treating infections during pregnancies seriously for a healthy baby. |
| **Outcome 1.1: Increased number of pregnant women attending frequent check-ups to treat infections** | Number of pregnant women who attend check-ups for infections | RCH report<br>Outpatient register |  |
| **Output 1.1.1 Catchment Areas are educated on the need to get checked up when they get infections during pregnancy to protect the life of their babies** | Number of people educated | Health promotion report<br>Outreach report | Catchment Areas are available for health education outreach |
| **Activities**<br>**1.1.1.1: Train pool of reproductive health educators**<br>**1.1.1.2: Develop outreach plan for health educators**<br>**1.1.1.3: Carry out reproductive health education outreach in Catchment Areas.** | Number of trained health educators<br>Number of outreaches conducted | Outreach report | Catchment Areas are available for health education outreach |
| **Objective 2: Educate women on healthy lifestyle for safe pregnancy** | Number of macerated stillbirth due to unhealthy lifestyle recorded | Reproductive and Child Health report (RCH)<br>Consulting room register | Catchment Areas are available for health education outreach |
| **Outcome 1.1: Increased number of pregnant women leading healthy lifestyle** | Number outreach conducted | RCH report<br>Outpatient register |  |
| **Output 1.1.1 Catchment Areas are educated on the need to lead healthy lifestyles for the health of their babies** | Number of people educated | Health promotion report<br>Outreach report | Catchment Areas are available for health education outreach |
| **Activities**<br>**1.1.1.1: Train pool of health educators**<br>**1.1.1.2: Develop outreach plan for health educators** | Number of outreaches conducted | Outreach report | Catchment Areas are available for health education outreach |
| 1.1.1.3: Carry out healthy lifestyle education for Catchment Areas. |  |  |  |
| Objective 3: Educate women especially pregnant women and their husbands on psychological and emotional stress management | Number of pregnant women who had macerated stillbirth due to stress | Reproductive and Child Health report (RCH)<br>Consulting room register | Catchment Areas are available for health education outreach |
| Outcome 1.1: Increased number of pregnant women managing stress effectively | Number outreach conducted | Counselling room registers |  |
| Output 1.1.1 Catchment Areas are educated on the need to effectively manage stress levels during pregnancy<br>Husbands are educated on how to support their wives during pregnancy. | Number of people educated | Health promotion report<br>Outreach report | Catchment Areas are available for health education outreach |
| **Activities**<br>**1.1.1.1: organize stress management exercises for pregnant women in the catchment areas**<br>**1.1.1.2: Recruit a group of counsellors who will speak to the targeted population**<br>**1.1.1.3: Develop an outreach plan Catchment Areas.** | Number of outreaches conducted | Outreach report | Catchment Areas are available for health education outreach |
| **Objective 4: Make mobile antenatal and postnatal services available to the catchment areas of Volta Region.**<br>**Pool resources from grants, donations and tokens to furnish maternity wards** | Number of mobile antenatal services organized.<br>Maternity ward is refurnished with new supplies and equipment | Mobile Outreach Report | Resources for mobile outreach available. |
| **Outcome 3.1: Increased number of antenatal care visit**<br>**Outcome 3.2: refurnish maternity wards** | Number of pregnant women who visit antenatal care | Internal RCH Report | Catchment Area mothers make use of mobile antenatal services |
| **Output 3.3: Catchment Area provided with mobile antenatal services** | Number of frontline ANC attendant members with requisite skills | Mobile Outreach Report |  |
| **Activities**<br>**3.1.1.1: Get funds for mobile services and refurnishing of maternity ward**<br>**3.1.1.2: Get mobile service team on board**<br>**3.1.2.3: Procure supplies and equipment for training**<br>**3.1.2.4: Conduct training for mobile service Team.** | Number of mobile antenatal services organized. | Mobile Outreach Report | Resources for mobile outreach available.<br>Long distance to facility for ANC service no more a barrier |

*Source: QI 2022 project*

### Key Stakeholders Identified

Stakeholders are individuals that have interest in a company or project of which they can either affect or be affected by a business.

- Heath care professionals
- Project sponsors
- Assembly man
- Community Chiefs
- Ghana health service

### Stakeholder Communication Plan

| AUDIENCE | MESSAGE | MEDIUM | SCHEDULE | RESPONSIBLE |
| --- | --- | --- | --- | --- |
| Health care professionals | Progress note | Formal | Quarterly | Group |
|  | Status report | paper-based |  | Leader/Project |
|  | Budget report | report<br>Email |  | Manager |
| Project sponsors | Progress note | Formal | Quarterly | Group |
|  | Status report | paper-based |  | Leader/Project |
|  | Budget report | report<br>Email and<br>calls |  | Manager |
| Assembly Man | Progress note | Formal | Quarterly | Group |
|  | Status report | paper-based |  | Leader/Project |
|  | Budget report | report<br>Email |  | Manager |

### Project Schedule

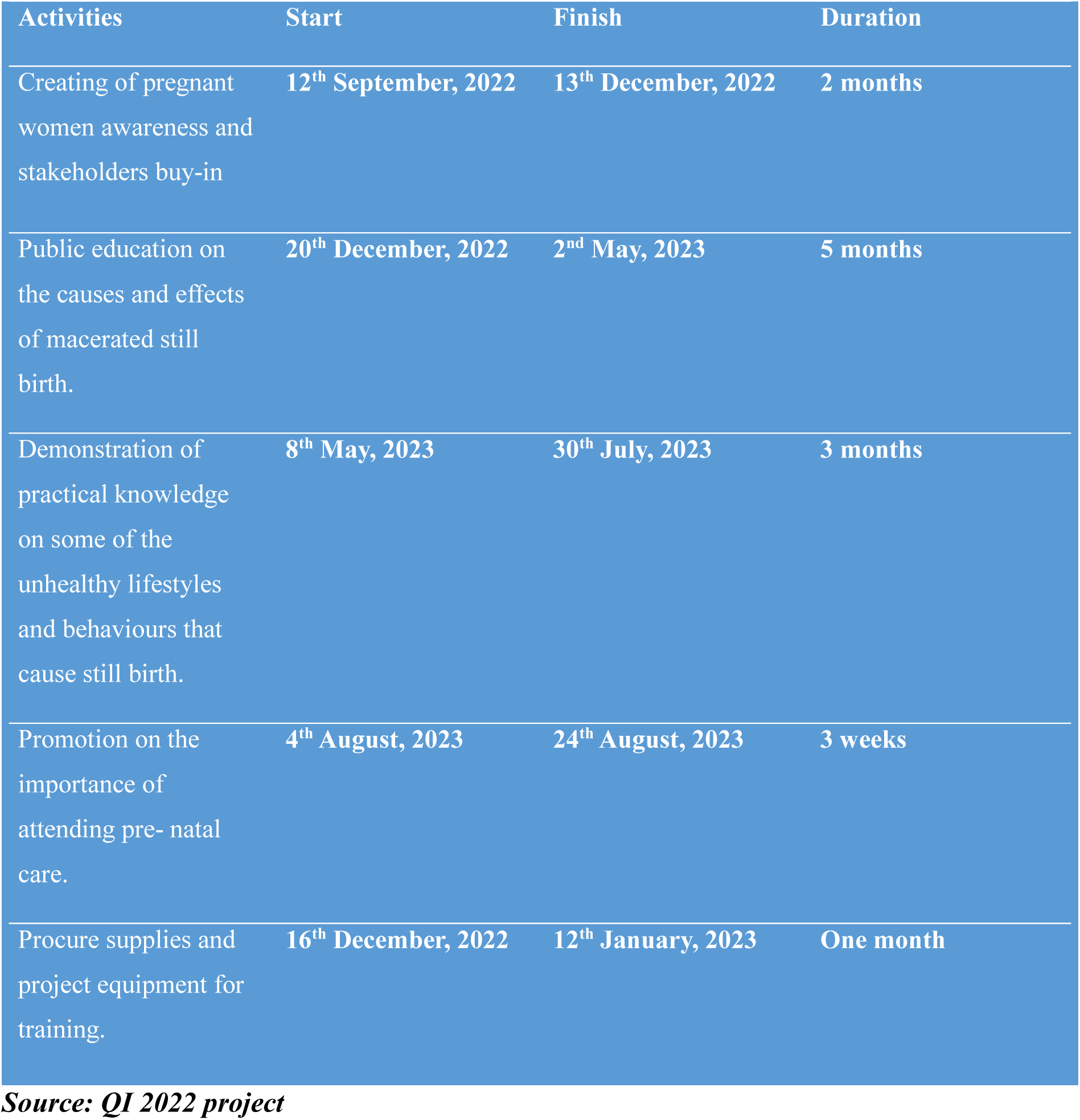

### Gantt Chart

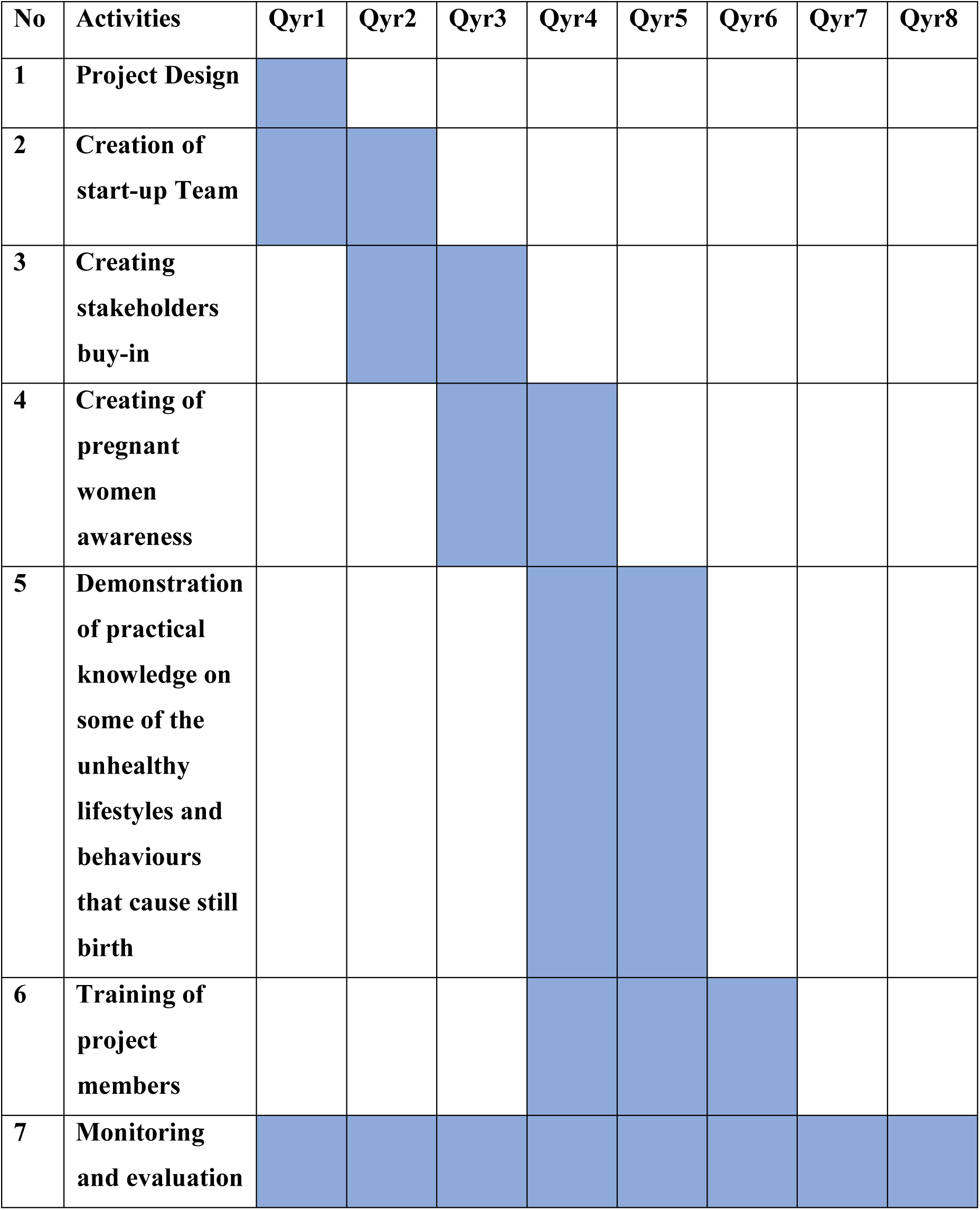
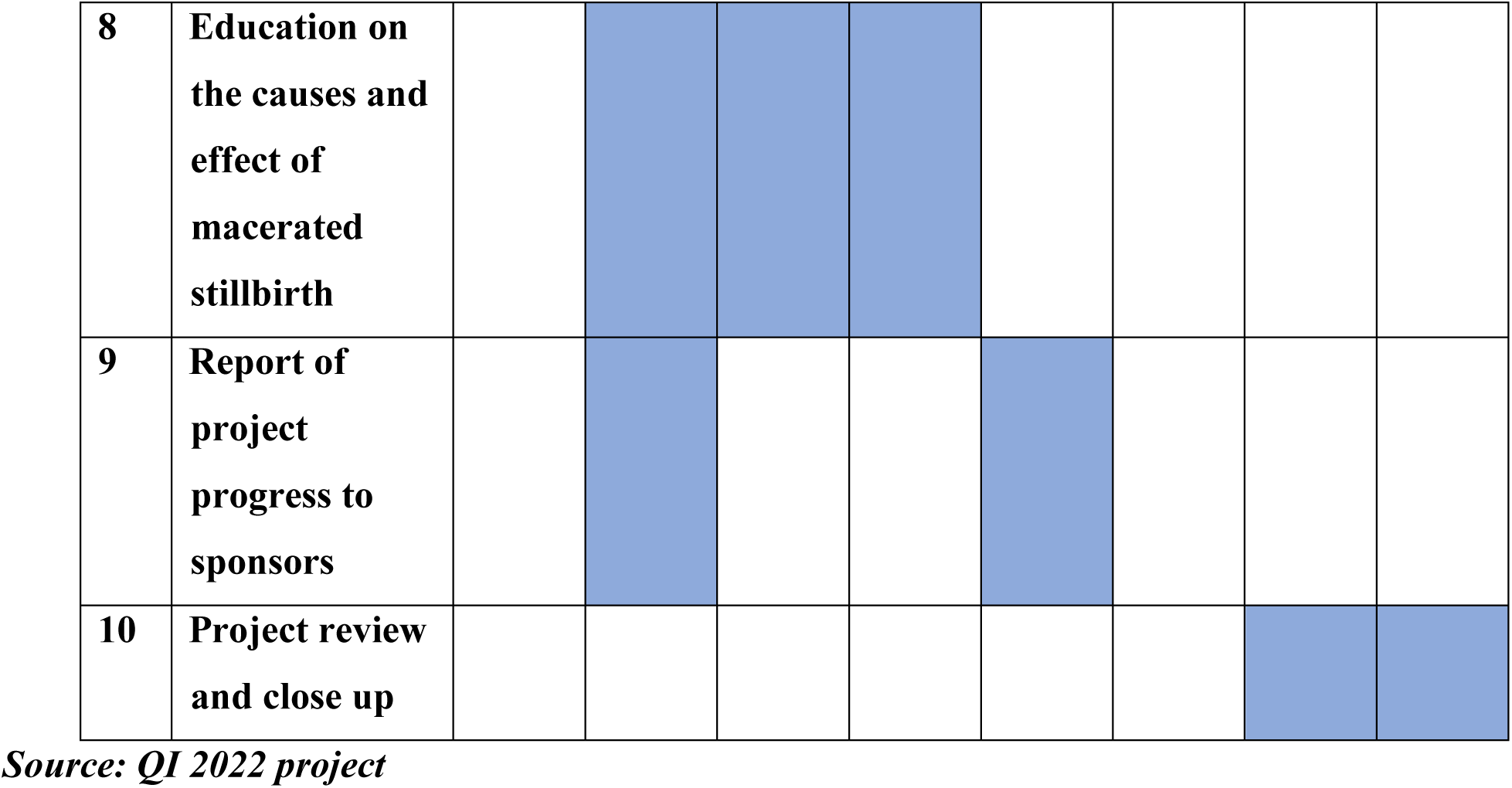

### Work Breakdown Structure

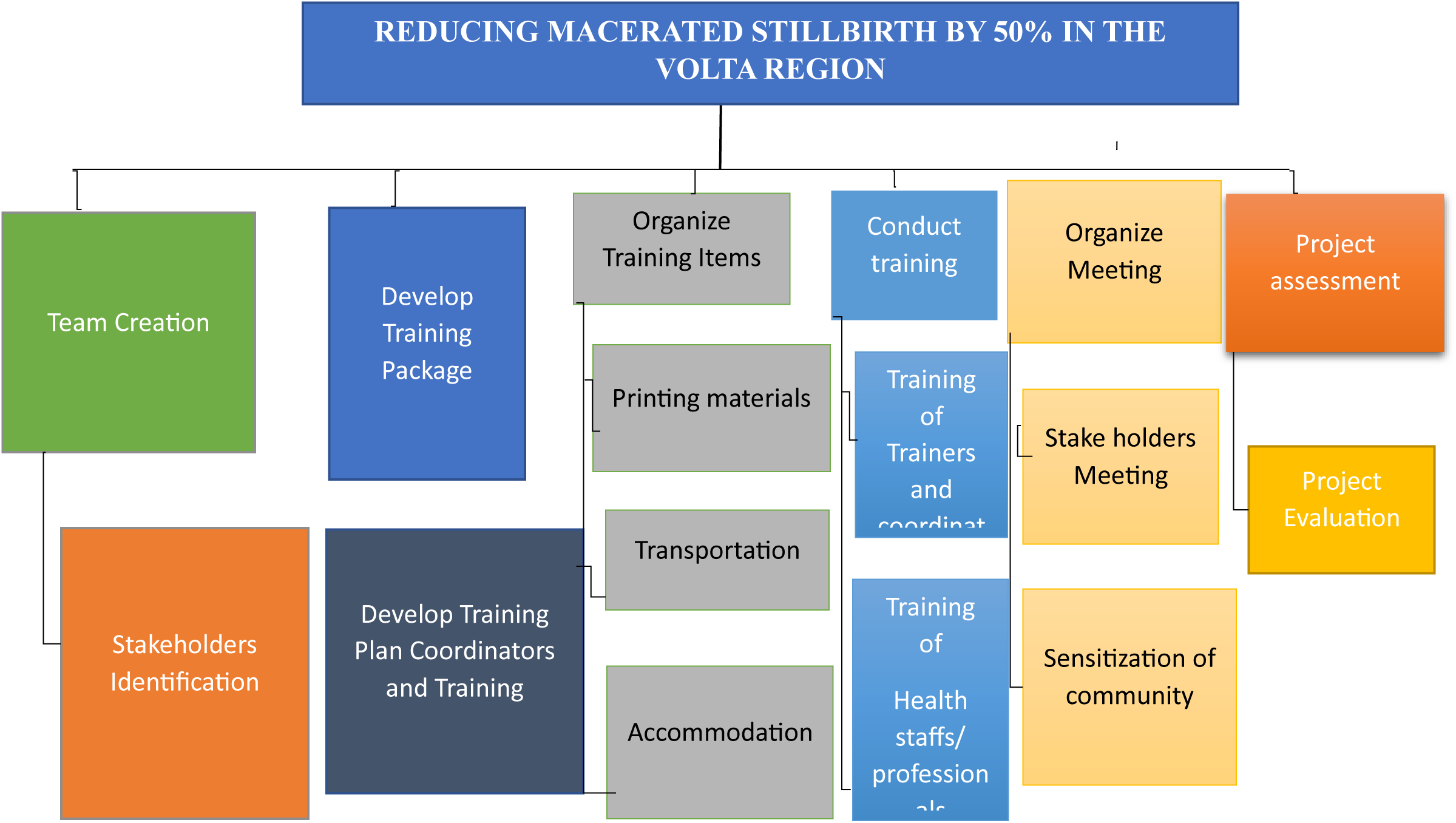

### Project Roles and Responsibilities

In a project, defining roles and responsibilities shows the levels of authority and ensures the smooth running of the project. Each member contributes effectively to the realization of the project. The table below shows group members roles assigned in achieving the aim of this project.

| MEMBER | ROLE | RESPONSIBILITIES |
| --- | --- | --- |
| GIDEON DANSO | PROJECT MANAGER | Ensures that the project is completed within the specified time frame and budget while meeting the project's goals. |
| GIDEON DANSO | RESOURCE MANAGER | Manages project-specific resource pools. Assigning the right people to the right project at the right time is your responsibility. |
| GIDEON DANSO | PROJECT SECRETARY | In charge of keeping track of all meetings and activities. |
| DR RICHARD OKYERE BOADU | MONITORING AND EVALUATION OFFICER | The monitoring and evaluation plan for the project is developed and implemented by this person. Coordinates systems for collecting and analysing data on the project's inputs, outputs, outcomes, and impact. |
| DR RICHARD OKYERE BOADU | DATA COORDINATOR | Responsible for the overall data management of a project |

*Source: QI 2022 project*

### Project Resources

In every project it is necessary to look at your resources both financially and humanly. The table below the list of resources we used for the accomplishment of our project within the 2 years in the Volta Region.

| RESOURCE TYPE | RESOURCE ITEM | QUANTITY/AMOUNT |
| --- | --- | --- |
| HUMAN RESOURCE | Midwives | 3 |
|  | Community Health Nurses | 12 |
|  | Gynecologists | 3 |
|  | Doctors | 6 |
|  | Health Information Officer | 2 |
| FINANCIAL | Budget | GHC 100,000.00 |
|  | Refurnish |  |
|  | Accommodation |  |

*Source: QI 2022 project*

### Project Budget Summary

| Budget line | Description | Unit Price (GHC) | Quantity | Total (GHC) |
| --- | --- | --- | --- | --- |
| Resource Materials for Training | Printing of IEC materials | 25.00 | 4000 | 1,000.00 |
|  | Printing of training Materials | 5.00 | 300 | 1,500.00 |
|  | White board Markers | 40.00 | 4 | 160.00 |
|  | Flip Chart | 60.00 | 30 | 1,00.00 |
|  | Pens | 55.00 | 2 | 120.00 |
|  | File Bag | 20.00 | 50 | 1,000.00 |
|  | Note Pads | 5.00 | 200 | 1,000.00 |
| Sub Total |  |  |  | **15,580.00** |
| Travel & Accommodation | Transport fare | 500.00 | 20 | 10,000.00 |
|  | Rent | 1,000.00 | 240 | 240,000.00 |
| Sub Total |  |  |  | **2,160.00** |
| Refurnishing | Labourers | 150.00 | 20 (10 days) | 30,000.00 |
|  | Materials | 100,000.00 | - | 100,000.00 |
| Sub Total |  |  |  | **10,000.00** |
| Community and Local Stakeholders Sensitisation Meetings | Venue Hiring | 1,000.00 | 4 | 4,000.00 |
|  | Drinks | 5.32 | 200 | 1,064.00 |
|  | Snacks | 60.00 | 200 | 12,000.00 |
|  | Fuel | 10.00 | 30 | 300.00 |
| Sub Total |  |  |  | **37,000** |
| Grand Total |  |  |  | **100,000.00** |

Source: QI 2022 project

## PHASE 2 (DO)

### Using the PDSA Quality Improvement Tool

PDSA is a model for improvement tool. It is a simple yet effective tool for implementing changes for improvement. It is the framework that was used to implement the ideas and solutions generated to help reduce the stillbirth rate at marry Lucy Hospital. It consists of two parts; a thinking part and a doing part. Using the PDSA QI tool, we set our aims, define measurements, find promising ideas for change, and test those ideas in real work.

What are we trying to accomplish (Goals and Objectives)?

### Main Aim

Reduce macerated stillbirth rate by 50% at the maternity unit in the eight (8) selected facilities in the Volta Region in 2 years’ time.

### Objectives

- Providing mobile services to ANC clients (pregnant women). services.
- Educating ANC clients (pregnant women) on Reproductive Health leading to safe delivery.
- Educating women (pregnant women) on stress management
- Unhealthy lifestyle of pregnant women

**Measurement** [How will we know that a change is an improvement]

Outcome measures:

Macerated still Birth Rate in the 8 selected facilities is reduced by 70% in 2 years’ time.

Process measures:

- Number of health education outreaches conducted
- Number of mobile antenatal services conducted
- Number of equipment provided at ANC room

In order to achieve our main objectives for this program we;

#### 1. Engaged patients in health facility and their families in QI program

Talk with a few patients on a daily basis.

Put patients and family members on the QI team.

#### 2. Make the chief financial officer a quality champion

We ensured financial officers understand the link between improving quality and decreasing cost. Let financial officers know they can encourage investment in QI.

#### 3. Engaged physicians

- Discover a common purpose: reducing mortality and morbidity.
- Identify and activate champions.
- Involve physicians from the beginning.
- Choose messengers and messages carefully.
- Value the physicians’ time.
- Communicate often and frankly.

#### 4. Build capacity in improvement

Ensure our group members have the knowledge and skills to teach quality and safety improvement.

### PDSA Cycle for the First Objective

A PDSA Cycle for the third objective: Make mobile antenatal services available for the pregnant women who visit the eight (8) selected facilities in the Volta Region.

**Figure 9.**
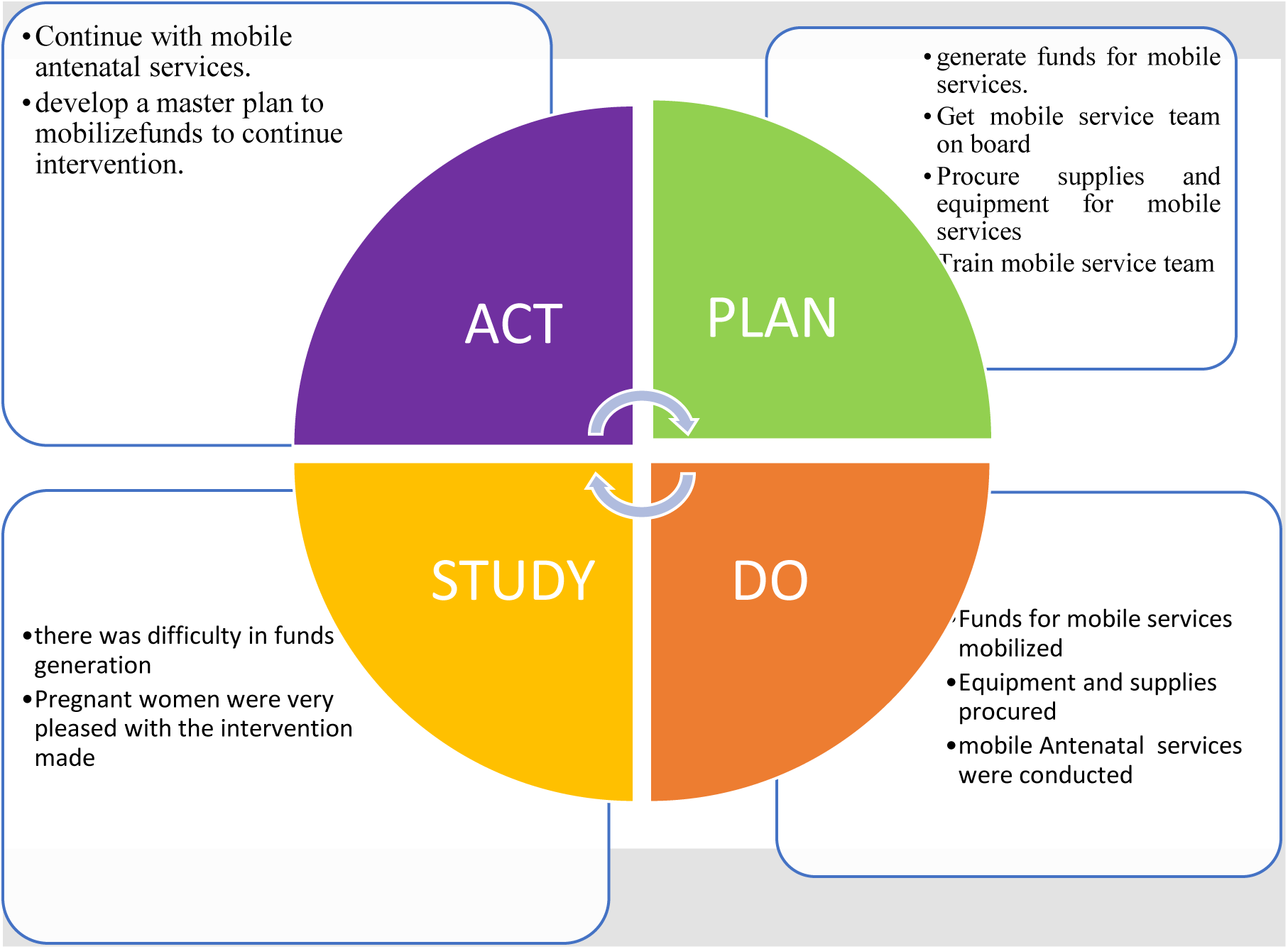

### Detailed PDSA for Figure 1. 9

#### PLAN

Generate funds for mobile services.

Get mobile service team on board

Procure supplies and equipment for mobile services

Train mobile service team

#### DO

Funds for mobile services successfully mobilized

Procured equipment and supplies

Mobile antenatal services were conducted

#### STUDY

There was difficulty in generating of funds

Pregnant women were very pleased with the intervention made.

#### ACT

Continue with mobile antenatal services.

Develop a master plan to mobilize funds to continue intervention.

### PDSA Life Cycle for Second Objective

PDSA cycle on conducting a reproductive education program for ANC clients (pregnant women) in eight selected facilities in the Volta Region.

**Figure 2.0.**
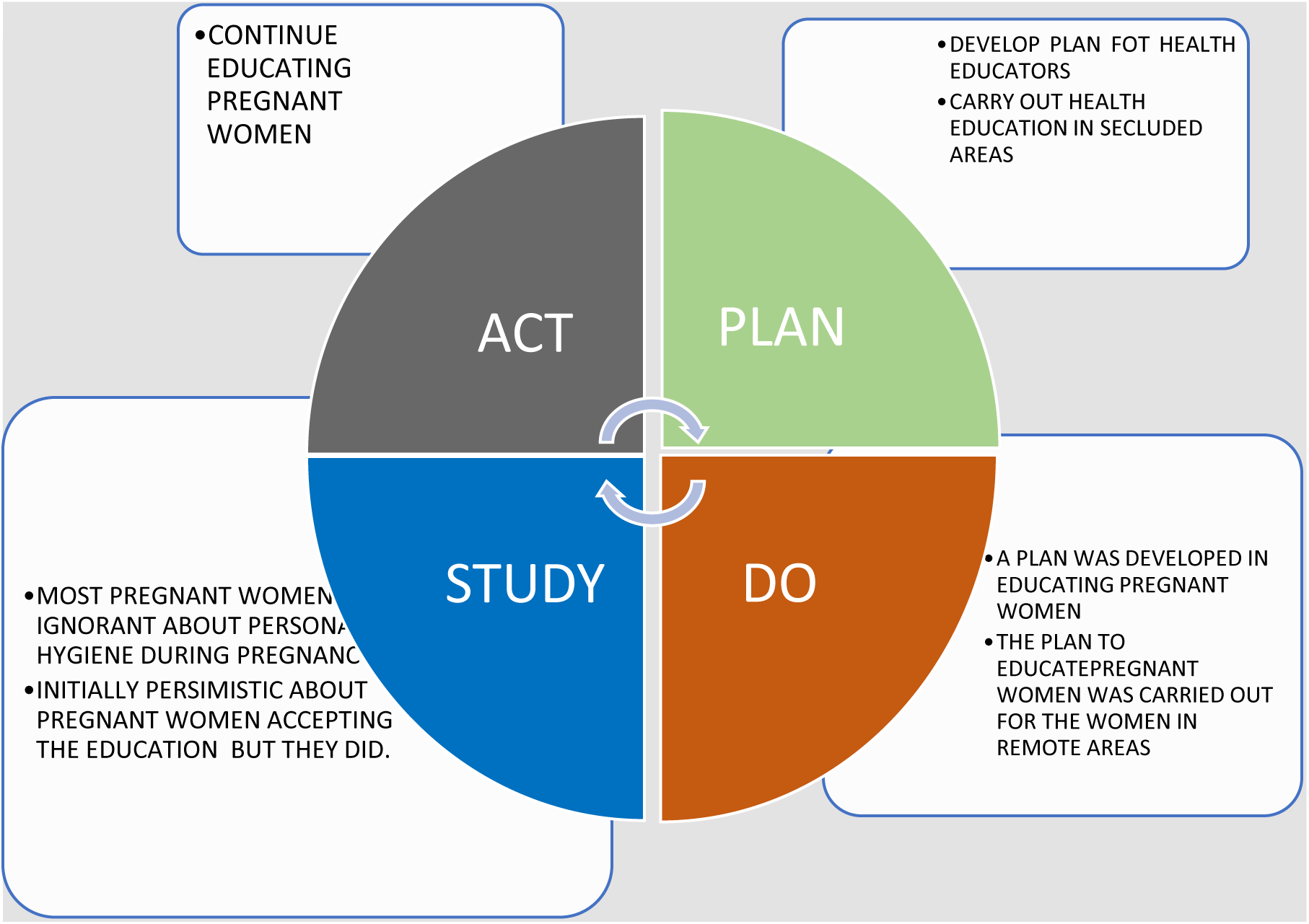

### Detailed PDSA for Figure 2.0

#### PLAN

Develop a plan for health educators

Carry out health education in secluded areas

#### DO

A strategy was developed in educating pregnant women

The plan to educate pregnant women was carried out for women in remote areas

#### STUDY

Most pregnant women were ignorant about personal hygiene during pregnancy and exercises Initially pessimistic about pregnant women accepting the education but they did.

#### ACT

Continue educating pregnant women

### PDSA Cycle for the Third Objective

A PDSA Cycle for the third objective: Educating women (pregnant women) who visit the eight (8) selected facilities in the Volta region on psychological and emotional stress management

**Figure 2.1.**
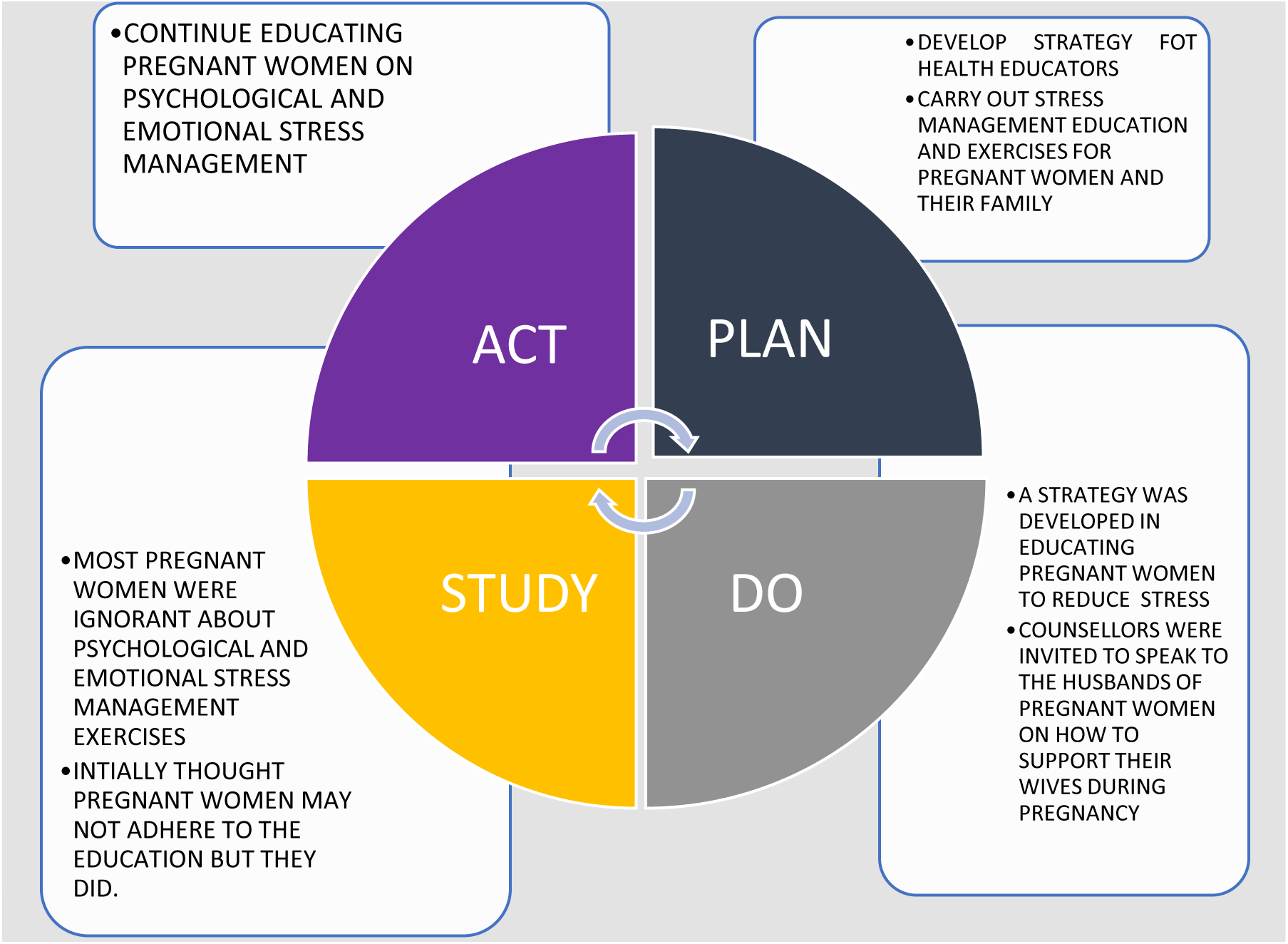

### Detailed PDSA for Figure 2.1

#### PLAN

Develop strategy for health educators

Carry out stress management education and exercises for pregnant women and their families

#### DO

A strategy was developed in educating pregnant women on how to reduce emotional and psychological stress

Counsellors were invited to speak to the husbands of pregnant women on how to support their wives during pregnancy period.

#### STUDY

Most of the pregnant women were ignorant about psychological and emotional stress management exercises.

Initially assumed that pregnant women will not accept the education but they did.

#### ACT

Continue educating pregnant women on psychological and emotional stress.

### PDSA Cycle for Fourth Objective

PDSA cycle unhealthy lifestyle of pregnant women in the Volta Region

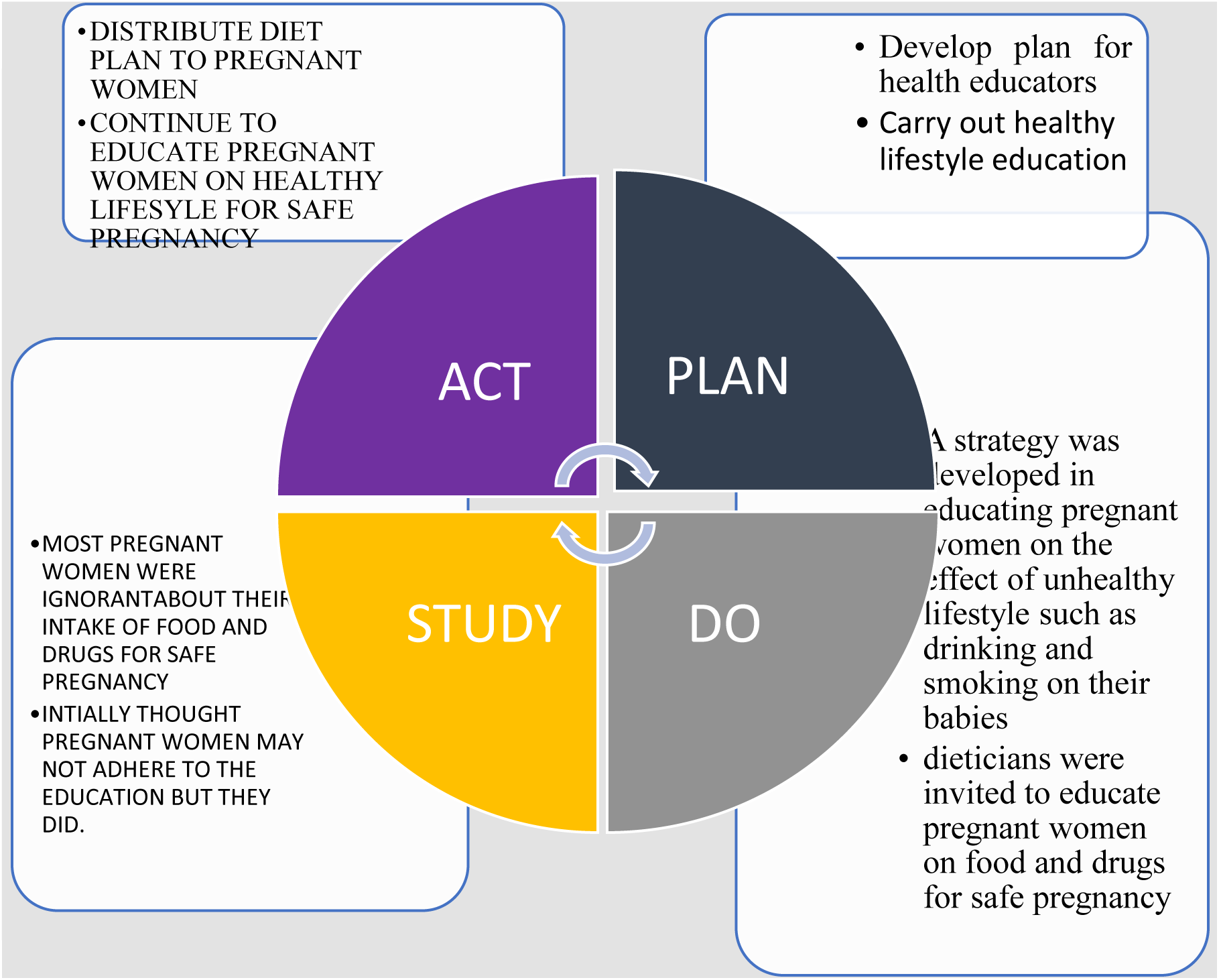

### Detailed PDSA for Figure 2,2

#### PLAN

Develop plan for health educators

Carry out healthy lifestyle education for pregnant women and their families

#### DO

A strategy was developed in educating pregnant women on the effect of unhealthy lifestyle such as drinking and smoking on their babies

Dieticians were invited to speak to pregnant women about diets and drugs needed for heathy pregnancy

#### STUDY

Most of the pregnant women were ignorant about their intake of food and drugs for safe pregnancy Initially assumed that pregnant women will not adhere the education but they did.

#### ACT

Distribute diet plan to pregnant women

Continue to educate pregnant women on healthy lifestyles leading to safe pregnancy

## PHASE 3: (STUDY)

This chapter focuses on risks identified, lessons learnt, risk monitoring and control.

### Risk Identified

Risk is the probability that an uncertainty will occur. These uncertainties have the potential to alter at least one or more of our objectives in a negative or positive way. In regard to this project, we put ahead the following risk;

- Risk of price increment on commodities (Inflation) and this will affect the project budget,
- Risk of local stakeholder participation and
- Risk associated with climatic conditions.

**Table 10:** Shows risks that we encountered during our project.

| IMPACT |  |  |  |  |  |  |
| --- | --- | --- | --- | --- | --- | --- |
| Probability |  | Insignificant | Marginal | Moderate | Critical | Catastrophic |
| Definitely |  |  |  |  | INFLATION |  |
| Likely |  |  |  |  |  | STAKE HOLDER<br>PARTICIPATION |
| Occasionally |  |  |  | CLIMATE<br>CHANGE |  |  |
| Seldom |  |  |  |  |  |  |

### Risk Monitoring Control

| Activity | Identified Risk(S) | Description | Impact Intensity | Mitigation Strategy | Contingency Plan |
| --- | --- | --- | --- | --- | --- |
| **Procurement of Resource Materials and Maintenance** | Inflation | Increment of items over time which will go against our budget | Critical | Procure all needed resource materials that will not expired over time, (e.g. fuel, printing materials) | Supplementary Budget (Miscellaneous) |
| **Stakeholder Engagement** | Low stakeholder participation | Stakeholders not having or losing interest in the project | Catastrophic | Organize monthly Stakeholders meeting to keep them updated on the project level |  |
| **Refurnishing of ANC unit** | Climatic change | Heavy Rainfall will cause distraction | Moderate | Renovation should be done in non-rainy season |  |

### Change Agent

In order to achieve this there is a need to select few staff at the facility to serve as change agents. Change agents are the ambassadors who can smooth the road by helping staff understand the benefits of this program. Barriers always occur during a change process; thus, the change agent needs to be alert and ready to help the staff work through the problems. The change agent also will work with management to assure needed resources are available and the required approvals are obtained.

We therefore determined what modifications should be made, mitigated high level risk and decided to plan our next test.

### Spread Framework

Spread help organizations build on processes that originate at the local level. Organizations need to be prepared to address external and internal factor that can affect spread

An effective spread plan involves

- Communication among the team
- Collaboration with other units
- Accounting for variability
- Starting with the highly adoptive group

### Developing a Plan for Spread

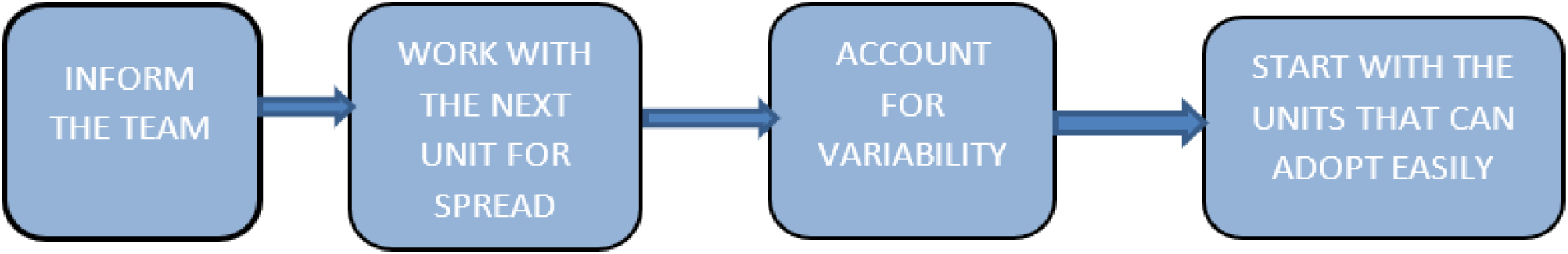

### Spread Framework Diagram

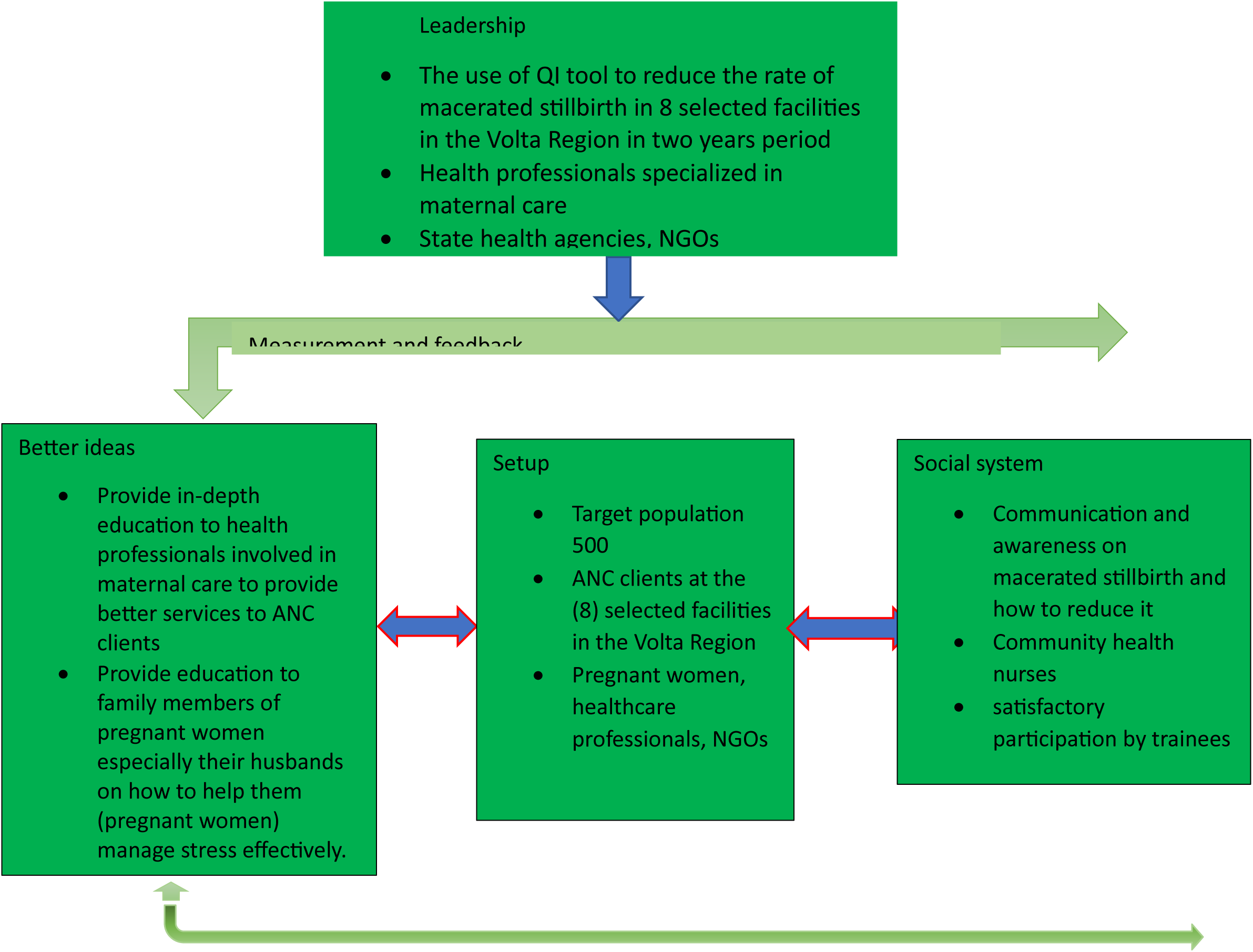

## PHASE 4 (ACT)

### Introduction

Implementing any change requires thoughtful planning. People resisting change is a common experience. Implementation of an intervention can be done in variety of ways, including adapting or developing policies and procedures, communication of the standards, conducting training programs, coaching/mentoring, providing supportive supervision and engaging the community to play their part in improving the quality of maternal and newborn health care. Prior to implementing these strategies, steps must be taken to prepare the health facility to make the changes needed to improve the quality of care and services. The intervention implemented in chapter three above, was to help reduce the rate macerated stillbirth by 50% at the selected hospitals in the Volta Region, the success of the intervention will reduce the mean from the initial 62.8 to about 31.4. After the intervention, we were able to reduce it by additional 10% bring the mean down from 31.4 to 21.4. We therefore decided as a group to adapt to this intervention as it will really help reduce macerated stillbirth to a really significant level and if possible, extends this program to the whole Volta Region. People are more likely to accept change if they see the need for the intervention and this result means that the people of the Volta Region saw a need for this intervention and hence accepted it.

### The graph below shows the results for the rate of macerated still birth before and after the intervention

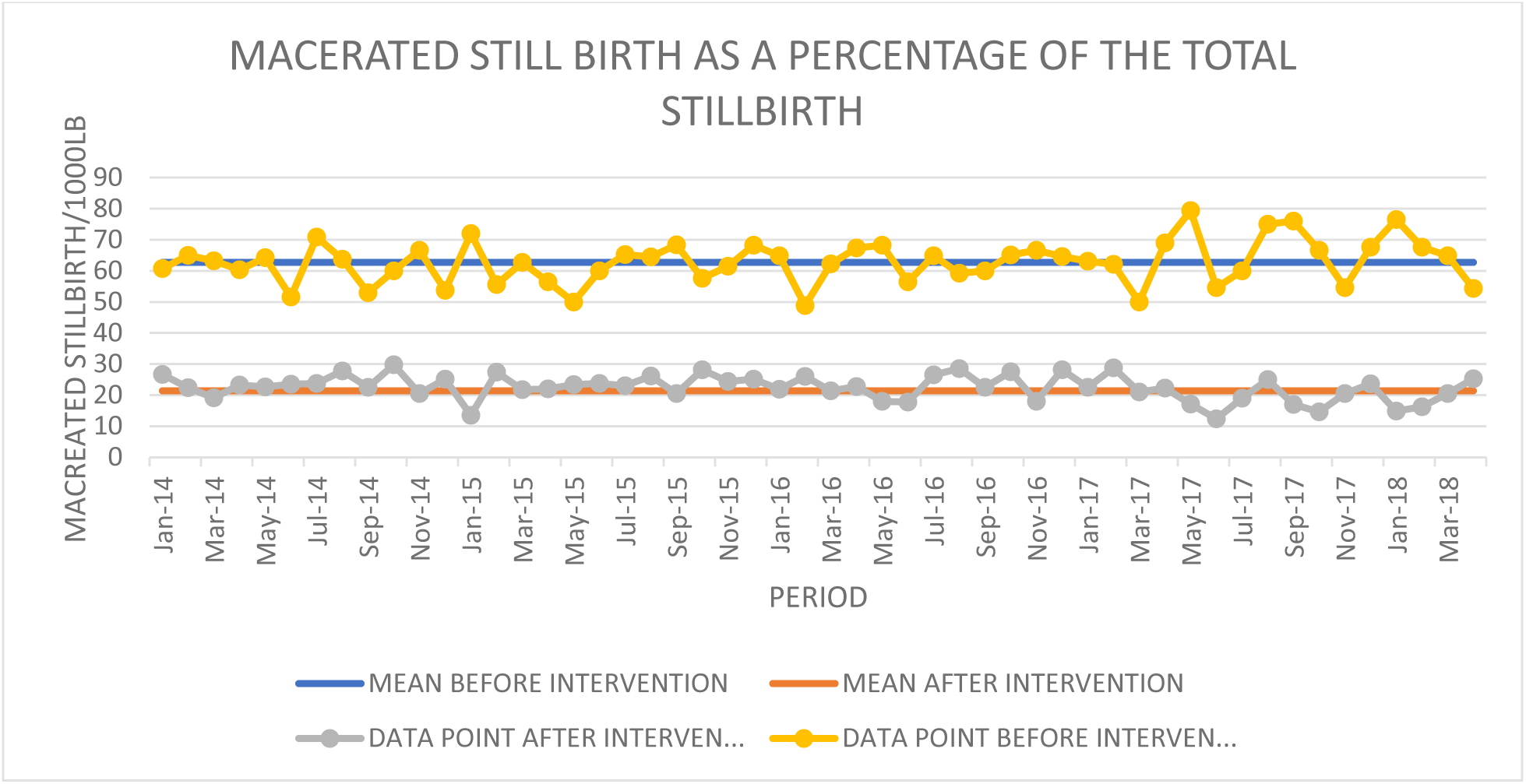

### Recommendations

- There should be regular education for ANC clients and relatives on Pregnancy care
- There should be a regular home visit to see how client are doing on Pregnancy care
- There should constant awareness on the effect of psychological and emotional stress on pregnant women.
- All stakeholders should be engaged in the quarterly Health Committee Meetings.

### Lessons Learnt

- Most healthcare professionals still need to be train on how to educate mothers who are unaware of how to live a healthy lifestyle for safe pregnancy.
- Most pregnant women were ignorant about their intake of food and drugs
- Initially thought pregnant women may not adhere to the education but they did.
- Getting funds were very difficult
- Pregnant women and mothers were very pleased with the intervention made

### Conclusion

In conclusion the use of the PDSA model helped reduce the rate of macerated still birth from 62.8per 1000 live births to 21.4 per 1000 live births. Implementing any change requires thoughtful planning. People resisting change is a common experience. Implementation of an intervention can be done in variety of ways, including adapting or developing policies and procedures, communication of the standards, engaging the community to play their part in improving the quality of maternal and newborn health care. A few families and pregnant women are still adamant and were not ready to change and this accounts to the reason why we were not able to reduce the rate of macerated stillbirth to the national target of 12/1000 live births or below. For effectiveness of the project a monthly and quarterly evaluation was done for do a good job. There is a room for improvement.

## Data Availability

All data produced are available at https://www.researchgate.net/publication/385354566_Neonatal_Health_Mortality_Rate_Data

## Key Definitions

**Stillbirth:** is defined by the Ghana Health Service as a baby born with no signs of life at or after twenty-eight (28) weeks of gestation.

**Fresh stillbirth:** total number of babies who died in the process of labour.

**Macerated stillbirth:** total number of babies who died in the uterus more than twelve hours before delivery with signs of maceration.

**Total stillbirth:** total number of babies who were delivered without signs of live.

**Total live birth:** total number of babies born alive.

**Neonatal death:** babies dying twenty-eight (28) days of life.

## Data Availability Statement

No new data were created or generated; however, the data analysed in this study is openly available and can be accessed via the link https://www.researchgate.net/publiation/385354566_Neonatal_Health_Mortality_Rate_Data

## Ethical Considerations

To ensure anonymity, no identifiable personal information such as name, telephone number, designation or position held by the study population were included the study. The data was use for only the primary purpose and authors were also thoroughly cited in the study.

